# Dynamic HIV risk differentiation among youth: Validation of a tool for prioritization of prevention in East Zimbabwe

**DOI:** 10.1101/2024.09.10.24312897

**Authors:** Louisa R Moorhouse, Simon Gregson, Jeffrey W Imai-Eaton, Justin Mayini, Tawanda Dadirai, Phyllis Magoge-Mandizvidza, Rufurwokuda Maswera, Simbarashe Mabaya, Rachel Baggaley, Daniel Low-Beer, Constance Nyamukapa, Shona Dalal

## Abstract

**Background:** Differentiating risk for HIV infection is important for providing focussed prevention options to individuals. We conducted a longitudinal study to validate a risk-differentiation tool for predicting HIV or HSV-2 acquisition among HIV-negative youth.

**Setting:** Population-based household survey in east Zimbabwe.

**Methods:** HIV and HSV-2 status and HIV behavioural risk factors were assessed in two surveys conducted 12 months apart among young people. Associations between risk-behaviours and combined HIV/HSV-2 incident infection were estimated using proportional hazards models. We calculated the sensitivity and specificity of risk-differentiation questions in predicting HIV/HSV-2 acquisition and quantified changes between surveys among low, medium, and high-risk categories.

**Results:** In total, 44 HIV/HSV-2 seroconversions were observed in 1812 person-years of follow up (2.43/100PY, 95%CI: 1.71-3.15); 50% of incident cases reported never having had sex at baseline. Risk of HIV/HSV-2 acquisition was higher for those reporting non-regular partners (women: HR=2.71, 95% CI:1.12-6.54, men: HR=1.37, 95%CI: 0.29-6.38) and those reporting having a partner with a sexually transmitted infection (STI) (HR=7.62 (1.22-47.51). Adding a question on non-regular partnerships increased tool sensitivity from 18.2% to 38.6%, and further to 77.3% when restricted to those who had ever had sex. Individual risk category increased for 28% of men and 17% of women over 12-months.

**Conclusion:** The refined risk differentiation tool identified a high proportion of youth at risk of HIV acquisition. Despite this, half of incident infections were among individuals who reported no prior sexual activity. The shifting patterns of risk behaviours underscore the need for dynamic prevention engagement strategies in high HIV prevalence or incidence settings.

## Introduction

Despite steady declines since 2000, UNAIDS estimated that 1.3 million people globally were newly infected with HIV in 2022^1^, well above targets to reduce new infections to fewer than 500,000 per year by 2020^1^. Multiple methods have been proven to prevent HIV acquisition, but access to and use of these methods varies. Identifying individuated risk of HIV acquisition is important to provide effective prevention options that meet individuals needs and manage programs. Risk-differentiation tools have been proposed to help to identify and refer people most in need of HIV prevention interventions^2^.

Risk-differentiation tools are used to determine one’s risk of acquiring HIV based on sociodemographic characteristics, circumstances, and sexual and partnership behaviour^3^. Priority populations with higher relative risk of HIV infection include adolescent girls and young women (AGYW) in eastern and southern Africa, and key populations including men who have sex with men^2^, those who engage in sex work or transactional sex and their partners, people who inject drugs^1^, trans and gender diverse people, and people in prison or other closed settings^4^. Risk-differentiation tools are developed empirically using available data from that population to identify predictors of HIV incidence. Although the World Health Organization (WHO) does not recommend risk tools to screen for HIV testing or exclude individuals seeking HIV prevention^5^ [CITE], in the context of declining overall incidence, it is important to focus the offer of prevention interventions that are intensive to deliver and adhere to, such as antiretroviral-based pre-exposure prophylaxis (PrEP)^6^. Moreover, risk assessment can support provider-client discussions about understanding individual need and demand for prevention services including among people accessing other routine health services^7^.

Studies have associated elevated risk of HIV acquisition with having multiple sexual partners^8^, non-regular and concurrent partners^9,10^, not knowing a partner’s HIV status^11^, young women having older male partners^12^, attending bars and beer halls^13^, presence of sexually transmitted infections (STIs)^14^ and sex within sero-discordant partnerships^15,16^. Existing risk screening tools are typically constructed with weights applied to specified risk factors calibrated in particular study populations and require adaptation for use in different populations or settings. The association of factors used as indicators of incidence can vary, and depends on the distribution of those with undiagnosed or untreated HIV infection and HIV prevalence within a population^2,17^.

HIV testing is an opportunity to identify and link HIV-negative persons to HIV prevention services^18^. Applying a tool at the point of an HIV-negative test result would cover a more general population than previous tools developed for specific at-risk populations^19^. With a focus on person-centred services, WHO proposed a series of questions to identify individual HIV risk level to guide referral to appropriate prevention packages including condom provision, VMMC, PrEP, community-based or key population programmes, depending on risk and personal preference^18. 19^Different from other risk-differentiation tools, the proposed approach does not use a threshold score to determine a binary prevention recommendation. Instead, HIV prevention interventions, and subsequent follow-up, can be offered depending on the level of risk determined by the tool or whether an individual requests prevention services. The aim was to better link the demand for prevention with the supply of services.

We measured the ability of behaviour questions to predict HIV or HSV-2 acquisition over 12-months among young people who participated in a general population household survey which informed the development and validation of a tool for WHO guidance on person-centred HIV strategic information [CITE]. Young people in Zimbabwe and across Southern Africa become sexually active in their mid-to late-teens and HIV incidence is high particularly among young women^1,20^. Sexually active young people could benefit from enhanced engagement with HIV prevention services^1^.

## Methods

### Study population and eligibility

This study was embedded in a two-round general population survey in eastern Zimbabwe^21^. A household census and questionnaire was conducted across 8 study sites, consisting of subsistence farming, agricultural estates, peri-urban and urban areas, between July 2018 and December 2019. Following household enumeration, females 15-24-years-old and males 15-29-years-old were invited to participate in an individual interview, home-based HIV testing and counselling (HTC), and collection of a dried blood spot sample (DBS). The interview included questions on sociodemographic characteristics, HIV knowledge, sexual risk, and use of HIV prevention methods. Informal confidential voting (ICV), whereby the respondent does not disclose their answers to the interviewer, was used in responses to sensitive questions^22^. HIV-negative young people at baseline were followed-up after 12-months and interviewed using a shortened questionnaire (Figure S1, Supplemental Digital Content).

This analysis was restricted to young people who completed the baseline and 12-month individual questionnaire, were HIV-negative at baseline and had an HIV test result at 12-month follow-up. HSV-2 status was ascertained for all HIV-negative young people at baseline and at 12-month follow-up for those who were not HSV-2 positive at baseline (Figure S1, Supplemental Digital Content).

This study was approved by the Medical Research Council of Zimbabwe, the Imperial College Research Ethics Committee (17IC4160), and the WHO Ethics Review Committee. Written consent was obtained from all adult participants. Assent was obtained for participants aged under 18 years alongside written informed consent from a parent or guardian.

### Risk-differentiation tool development and classification

Questions on individual and sexual partner risk-behaviours potentially associated with HIV incidence were identified through a literature review. Questions relevant across populations and HIV epidemic contexts were combined into a tool for validation. These questions covered self and partner risk-perception, having a known HIV-positive partner, having multiple sex partners in the past year, having a sexually transmitted infection in the past year, transactional sex, sex between men, and using injecting drugs. Several additional questions on sexual behaviour were part of the questionnaire administered at baseline and follow-up with the aim of developing a tool to identify individuals for referral to HIV prevention services. The draft tool and the equivalent questions are in Figure S2 (Supplemental Digital Content) and Figure S3 (Supplemental Digital Content). We defined risk behaviour as responding ‘yes’ to any of the risk behaviour questions and categorized high risk as reporting at least one of recent sex between men, recent non-regular partner or recent transactional sex; medium risk as reporting at least one of perceiving a high current or future risk of HIV infection, sex with multiple partners in the last 12 months, spouse/partner having other partners, symptoms of sexually transmitted infections (STI) in the last 12 months or awareness that last sexual partner is HIV positive; and low risk as having started sex but no sexual risk behaviours. The shortened follow-up survey risk category measurements did not include data on self or partner STIs, or sex between men.

### HIV and HSV-2 laboratory methods

HTC was carried out using the Zimbabwe Ministry of Health rapid-testing algorithm^23^. DBS were collected from all consenting individuals completing the individual questionnaire. Where HTC was declined, consent was obtained to conduct the same testing using DBS at the Biomedical Research and Training Institute (BRTI) laboratory^31^ (Figure S4, Supplemental Digital Content).

Baseline HSV-2 testing was conducted on participants who were HIV-negative according to study testing and provided DBS and consent for laboratory testing. HSV-2 status was established from ELISA testing of DBS at the BRTI laboratory (Figure S4, Supplemental Digital Content). HSV-2 ELISA results were recorded as negative, equivocal, or positive. Samples which remained equivocal following retesting were recoded as negative^24^.

Combined HIV/HSV-2 incidence was the primary measure of risk over the 12-month follow-up period, against which the predictive power of the risk screening tool was measured. HIV and HSV-2 incidence were calculated among the study population who were HIV and HSV-2 negative at baseline respectively. Individuals who tested HIV-negative or HSV-2 negative at baseline were followed up, and anyone seroconverting from HIV-negative to HIV-positive or from HSV-2-negative to HSV-2-positive during the study follow-up time was classified as a combined risk seroconversion (Figure S6, Supplemental Digital Content).

### Statistical analysis

Sociodemographic characteristics and baseline risk characteristics of eligible individuals, stratified by sex, were described by sample proportions. Differences in proportions by sex were assessed using Chi-squared statistics. Socioeconomic status was measured using a household wealth index^25^. The precise date of HIV or HSV-2 infection in the inter-survey period was unknown, so seroconversion dates were randomly assigned thirty times and resulting incidence rates were combined using Rubin’s rules. The association of each risk behaviour with HIV/HSV-2 seroconversion was calculated using Cox proportional hazards models, adjusted for study-site and age. Sensitivity and specificity of the risk-differentiation tool to predict combined HIV/HSV-2 incidence were calculated among all participants and then restricted to individuals reporting to have started sex (sexual debut) at baseline. The change in sensitivity and specificity of the tool when adding questions was measured. Risk-behaviours during follow-up, reported at the 12-month survey, were described for individuals who seroconverted but met no criteria for baseline prevention referral. The proportions of individuals who changed risk categories between baseline and follow-up were examined using a Sankey diagram. The association of each risk behaviour reported at follow-up with HIV/HSV-2 seroconversion was assessed using logistic regression. Cluster robust confidence intervals, using village as the unit of clustering, were used throughout. Statistical analyses used STATA MP17 and Python.

## Results

At baseline, 1210 women 15-24-years and 1048 men 15-29-years participated in the survey (Table S1, Supplemental Digital Content). Follow-up questionnaires were completed by 81.5% (986/1210) of women and 65.8% (690/1048) of men; 98% of whom had HIV test results and 67% of whom had HSV-2 test results at baseline and follow up. 79% of participants took up HTC in the survey; 75% of those who declined testing self-reported being HIV-positive and on ART. Most participants were aged 15-19 years, had secondary education or higher, and had never married (Table 1). HIV prevalence was 3.0% (95% CI:2.1-4.2%) among women and 2.2% (95% CI:1.3-3.6%) among men. Among HIV-negative participants, HSV-2 prevalence was 21.7% (95% CI:19.0-24.6%) among women and 18.3% (95% CI:15.4-21.6%) among men.

**Table 1.** Baseline sociodemographic characteristics and HIV prevalence among adolescents and young adults completing 12-month follow up in eastern Zimbabwe.

|  | Women | Men | P-value of difference by sex |
| --- | --- | --- | --- |
|  | 15-24 years<br>N = 986<br>n (%) | 15-29 years<br>N = 690<br>n (%) |  |
| Age group |  |  |  |
| 15-19 years | 644 (65.3) | 374 (54.2) | <0.01 |
| 20-24 years | 342 (34.7) | 182 (26.4) |  |
| 25-29 years | 0 (0.0) | 134 (19.4) |  |
| Education |  |  |  |
| None/primary | 105 (10.7) | 51 (7.4) | 0.024 |
| Secondary/higher | 881 (89.4) | 639 (92.6) |  |
| Marital status |  |  |  |
| Never married | 616 (62.5) | 521 (75.5) | <0.01 |
| Currently married | 326 (33.1) | 159 (23.0) |  |
| Divorced/separated | 43 (4.4) | 10 (1.4) |  |
| Widowed | 1 (0.1) | 0 (0.0) |  |
| Socioeconomic status |  |  |  |
| Poorest | 108 (11.0) | 65 (9.4) | <0.01 |
| 2nd poorest | 397 (40.3) | 343 (49.7) |  |
| 3rd poorest | 261 (26.5) | 173 (25.1) |  |
| 4th poorest | 205 (20.8) | 101 (14.6) |  |
| Least poor | 15 (1.5) | 8 (1.2) |  |
| HIV status |  |  |  |
| Positive | 29 (3.0) | 15 (2.2) | 0.334 |
| Negative | 957 (97.0) | 675 (97.8) |  |
| Site type |  |  |  |
| Urban | 202 (20.5) | 119 (17.2) | <0.01 |
| Peri-urban | 266 (27.0) | 141 (20.4) |  |
| Estates | 225 (22.8) | 231 (33.5) |  |
| Rural | 293 (29.7) | 199 (28.8) |  |

### Baseline risk behaviour

At baseline, 42.2% of young women and 38.8% of young men reported ever having had sex (*Table 2*. High proportions of those who had debuted sexual activity reported having a non-regular partner (women: 30.0%; men: 67.2%) and HIV-positive women and men were more likely to report a spouse having other partners. More women reported recent STI symptoms and age-disparate relationships than men. Overall, 30% of participants were classified in the high-risk category, 7% in medium, and 4% in low risk, and the remaining 59% reported being not sexually active at baseline (no risk). When restricted to HIV-negative participants, HIV risk factors were more commonly reported by men than women, except for recent transactional sex. Among HIV-negative men, 1.5% reported sex with another man in the last 12-months. At baseline, 24% of sexually experienced participants reported using a condom at last sex; 41% of those in the higher-risk category reported using a condom at last sex compared to 8% of those in the low-risk category. Fewer than 1% of women had ever taken PrEP. 34% of men reported VMMC. 25% of HIV-negative women and 10% of HIV-negative men reported being previously sexually active, but not in the past 12 months (classified as not reporting any risk behaviours; Figure S7, Supplemental Digital Content).

**Table 2.** Baseline HIV risk behaviours by HIV status.

| Ages (years) | Female |  | Male |  |
| --- | --- | --- | --- | --- |
|  | 15-24 years |  | 15-29 years |  |
|  | % of HIV positive<br>(95% CI) | % of HIV negative<br>(95% CI) | % of HIV positive<br>(95% CI) | % of HIV negative<br>(95% CI) |
| N | 29 | 957 | 15 | 675 |
| Self-report to have ever had sex | 41.4 (23.5-61.1) | 42.4 (39.3-45.6) | 40.0 (16.3-67.6) | 38.8 (35.1-42.6) |
| Perceives a high chance of current HIV infection | 41.7 (15.2-72.3) | 1.2 (0.4-2.9) | 0.0 (0.0-45.9) | 0.7 (0.1-2.7) |
| Perceives a risk of HIV infection in next 12 months | 16.7 (2.1-48.4) | 0.7 (0.2-2.1) | 0.0 (0.0-45.9) | 3.1 (1.3-5.9) |
| Had sex with more than one partner in last 12 months | 0.0 (0-26.5) | 3.2 (1.7-5.4) | 33.3 (4.3-77.7) | 19.8 (15.2-25.2) |
| Had symptoms of sexually transmitted infection in last year | 16.7 (2.1-48.4) | 4.4 (2.6-6.9) | 0.0 (0.0-45.9) | 1.5 (0.4-3.9) |
| Ever involved in non-marital relationship where received anything in exchange for sex | 0.0 (0.0-26.5) | 3.4 (1.9-5.7) | 50.0 (11.8-88.2) | 14.5 (10.5-19.4) |
| Given or received money/goods/services in exchange for sex in last month | 0.0 (0.0-26.5) | 8.6 (6.1-11.8) | 16.7 (0.4-64.1) | 8.4 (5.3-12.4) |
| Among men: had sex with one or more man/men in last 12 months | - | - | 0.0 (0.0-45.9) | 1.5 (0.4-3.9) |
| Aware that last sexual partner is HIV positive | 33.3 (9.9-65.1) | 0.7 (0.2-2.1) | 0.0 (0.0-45.9) | 0.8 (0.1-2.7) |
| Spouse/partner has other partners | 16.7 (2.1-48.4) | 5.4 (3.4-8.1) | 16.7 (4.2-64.1) | 7.3 (4.4-11.1) |
| Regular partner has had a sexually transmitted infection in last 12 months | 0.0 (0-26.5) | 1.0 (0.3-2.5) | 0.0 (0.0-45.9) | 1.1 (0.2-3.3) |
| Had a non-regular partner in last 12 months | 41.2 (15.2-72.3) | 30.0 (25.6-34.8) | 50.0 (11.8-88.2) | 67.2 (61.1-72.8) |
| Attended a bar or beer hall in last 12 months | 0.0 (0.0-26.5) | 3.2 (1.7-5.4) | 33.3 (4.3-77.8) | 40.8 (34.8-47.1) |
| In age-disparate relationship of 10 or more years | 16.7 (0.4-64.1) | 0.9 (0.1-3.3) | - | - |
| Concurrent sexual partners | 0.0 (0.0-26.5) | 0.2 (0.00-0.13) | 0.0 (0.0-45.9) | 8.0 (5.0-12.0) |

*In this analysis, all risk behaviours restricted to those who have reported to have ever had sex*

### HIV/HSV-2 incidence and associations with self-reported risk behaviour

A combined 44 HIV or HSV-2 seroconversions were observed in 1812 person years (PY) of follow-up, corresponding to an incidence rate of 2.43/100PY (95% CI:1.71-3.15) (Table S2, Supplemental Digital Content). The combined incidence rate was 2.44/100PY (95% CI:1.50-3.52) in women and 2.40/100PY (95% CI:1.29-3.52) in men. There were five HIV seroconversions in 1812 PY, an HIV incidence rate of 0.28/100PY (95% CI:0.03-0.52) (females=3/1064PY (0.28/100PY); males=2/749PY (0.27/100PY)) and 39 HSV-2 seroconversions in 1132 PY giving an incidence rate of 3.45/100PY (95% CI:2.37-4.53) (females=23/645PY (3.57/100PY); males=16/487PY (3.29/100PY)). Half (22/44) of the incident cases of HIV and HSV-2 were among participants who reported never having had sex at baseline.

Among women, the sexual behaviours associated with HIV/HSV-2 seroconversion were having a non-regular sexual partner in the last 12-months (HR:2.71, 95% CI:1.12-6.54) or having a partner diagnosed with a STI in the past 12 months (HR:7.62 95% CI:1.22-47.51) (*Table 3)*. None were significant among men. Some risk-behaviours were weakly associated with seroconversion, including for women: reporting a spouse or regular partner having other partners (HR: 2.34, 95% CI:0.78-7.00) and having had symptoms of an STI in the past 12 months (HR: 3.41, 95% CI:0.75-15.50). There were no seroconversions among young people who perceived a future risk of HIV infection, were aware of having an HIV-positive partner, were men who have sex with other men, or who had concurrent sexual partners (*Table 3*).

**Table 3.**
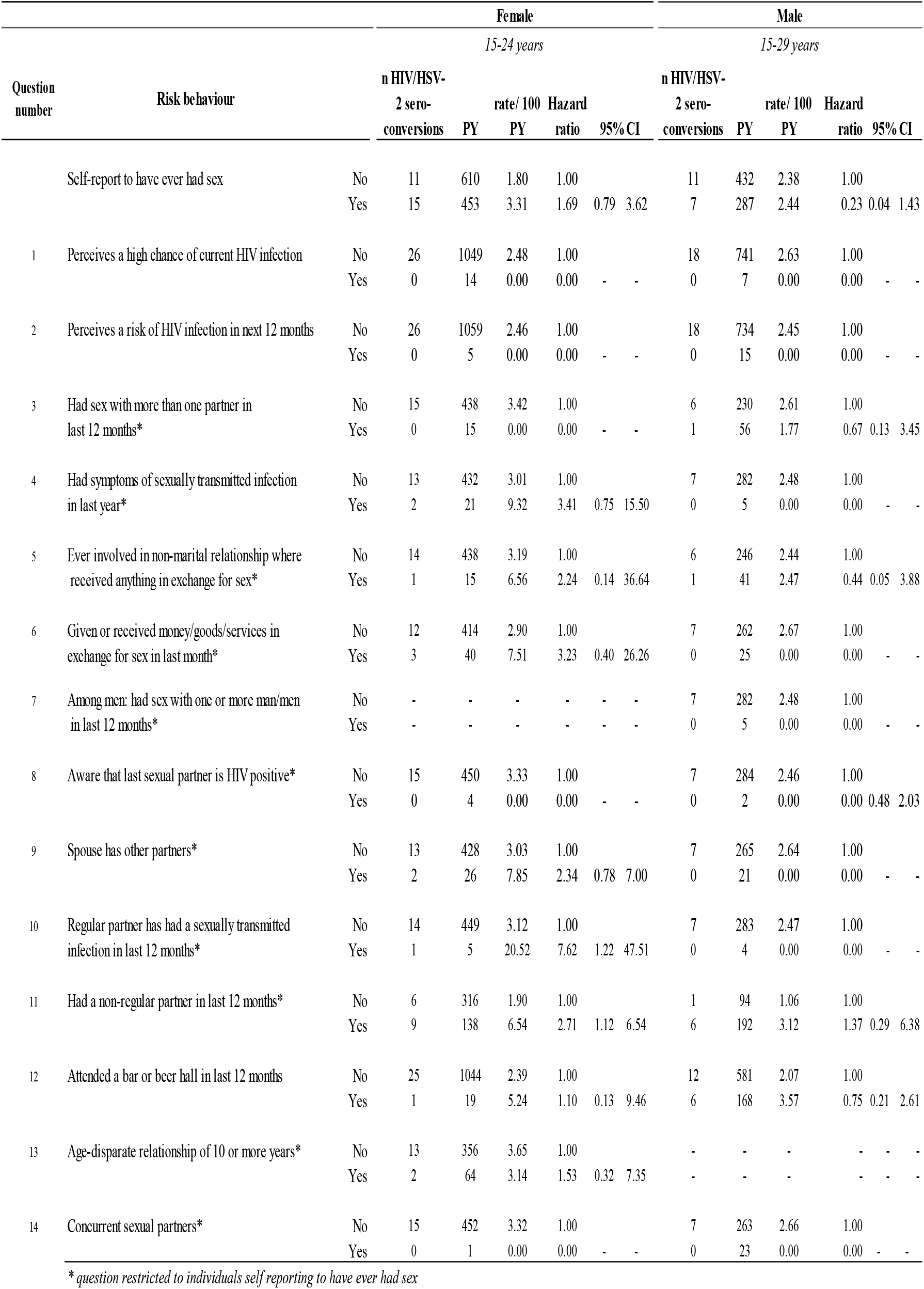
Associations of HIV risk-behaviours reported at baseline with combined HIV/HSV-2 incidence in baseline HIV/HSV-2-negative participants, stratified by sex and after adjusting for site and single year of age.

### Changes in HIV risk-behaviour and HIV/HSV-2 incidence over time

Sexual debut changed for 38% of men (157/413) and 14% of women (79/551) over the 12-month follow-up period (*Figure 1*). In this group, 10.3% of women and 26.6% of men moved from no sexual debut to medium- or higher-risk categories during the 12-month follow up period.

**Figure 1.**
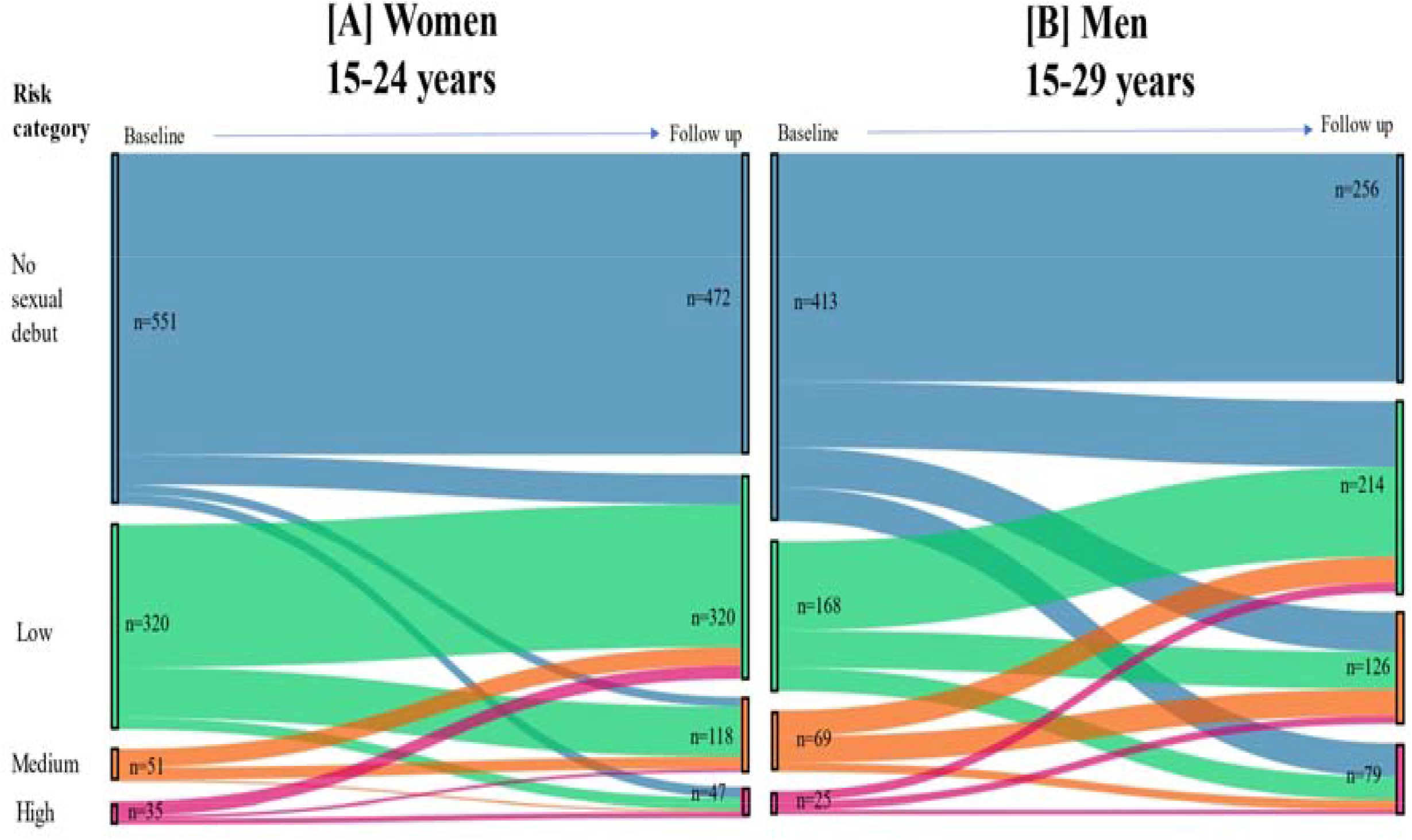
Changes in risk categories from baseline to 12-month follow-up among adolescents and young people testing HIV and HSV-2 negative at baseline in eastern Zimbabwe

Among individuals who had sexual debut at baseline, 31.3% of women and 39.7% of men in the low-risk category moved to medium- or high-risk categories. Conversely, 42.3% of men and 57.7% of women moved from a medium- or high-risk category at baseline to a low-risk category at follow-up. Overall, 28% of all men and 17% of women increased risk category from baseline to 12-month follow up.

Of the 36/44 HIV/HSV-2 incident cases observed among those not classified in medium- or higher-risk group at baseline, 16.7% (6/36) reported having multiple sexual partners in the last 12-months, 30.7% (1/36) reported having a non-regular sexual partner in the last 12-months, none reported having concurrent sexual partners, and 11.1% (4/36) reported recent transactional sex at 12-month follow-up. 5.5% (2/36) remained classified as low risk at 12-months.

### Risk tool sensitivity and specificity

Combining questions 1-10 (*Table 3)* into a tool would have identified 26.9% (7/26) of the women and 5.6% (1/18) of the men who subsequently seroconverted as being eligible for HIV prevention referral. This combination of questions had a sensitivity of 18.2% (8/44) and a specificity of 88.3% (1401/1586) to predict combined HIV/HSV-2 acquisition (*Table 3*). When restricted to those who reported sexual debut, sensitivity was 36.4% (8/22) and specificity was 73.5% (474/645). Adding a question on non-regular sexual partnerships (*Table 3 – Question 11)* to the tool increased the sensitivity to predict HIV/HSV-2 acquisition from 18.2% (8/44) to 38.6% (17/44) among all participants and from 36.4% (8/22) to 77.3% (17/22) among participants who had reached sexual debut at baseline (*Table 4*).

**Table 4.** Sensitivity and specificity of a risk-differentiation tool to detect combined HIV/HSV-2 incidence among young men and women in east Zimbabwe, 2018-2020.

|  | Sexually active at baseline |  |  | All at baseline |  |  |
| --- | --- | --- | --- | --- | --- | --- |
|  | All<br>N=686 | Female<br>N=418 | Male<br>N=268 | All<br>N=1676 | Female<br>N=986 | Male<br>N=690 |
| **Risk differentiation tool questions 1-10\*** |  |  |  |  |  |  |
| Sensitivity | 36.4% | 46.7% | 14.3% | 18.2% | 26.9% | 5.6% |
| Specificity | 73.5% | 79.8% | 63.8% | 88.3% | 90.2% | 85.6% |
| **Final risk differentiation tool for HIV prevention referral\*\*** |  |  |  |  |  |  |
| Sensitivity | 77.3% | 73.3% | 85.7% | 38.6% | 42.3% | 33.3% |
| Specificity | 51.8% | 61.1% | 28.3% | 78.1% | 82.4% | 71.9% |

*\*Perceives high chance of current or future HIV infection, sex with more than one partner in last 12 months, symptoms of STI in last 12 months, partner has symptoms of STI in last 12 months, ever or recent transactional sex, last sexual partner is HIV positive, spouse has other partners*
*\*\*as above with the addition of reporting a non-regular partner in last 12 months*

## Discussion

Understanding HIV risk and providing more focused prevention services is an ongoing need to reduce new HIV infections. Our results show that behavioural questions were associated with elevated risk of HIV/HSV-2 risk over 12-months among youth in eastern Zimbabwe. Combining these questions in a risk-differentiation tool gave a sensitivity of 39% and specificity of 78% among all participants, or 77% sensitivity and 52% specificity when restricted to sexually active participants. However, half of all incident infections occurred among participants who reported not being sexually active at baseline. Behaviour changed substantially during follow-up, underscoring the need for frequent offers of screening and of prevention services in settings with high background HIV prevalence or incidence.

Other studies have estimated risk differentiation tool sensitivities ranging from 30-90%, depending on the cut-off used^16,26,27^. Analysis of the VOICE risk-scoring tool in young women to decide PrEP eligibility found high sensitivity (above 80%) was only achieved with lower specificity (<50%) and by using a risk score threshold which meant most study participants were eligible for PrEP^26^. A systematic review of risk scoring for predicting HIV incidence concluded that the ability of risk scores to predict HIV incidence was only moderate^19^. They note that studies need to consider community-level HIV prevalence and the changing epidemiologic context in which tools are implemented. Among sexually active adult women, behaviours identified as consistently predictive of HIV acquisition were young age, non-cohabiting partnerships and presence of STIs^19^. Although STI status was included in the tool evaluated here, this was based on self-reported syndromic measures rather than laboratory confirmed status^19^.

In our study, 22 of 44 HIV/HSV-2 seroconversions occurred among those reporting no baseline sexual debut, reflecting a combination of rapid changes in sexual behaviour during a key period in the life-course, or under-reporting, which can be particularly problematic at young ages^28^. Over half of study participants who reported no baseline sexual debut may not have been referred for HIV prevention depending on how frequently the population was engaged. High proportions of those in the highest risk categories at baseline continued to report high risk sexual behaviours at 12-month follow-up. Although the proportions reporting high risk sexual behaviours at 12-month follow up were lowest among those in the lowest baseline risk category, risk behaviours changed for a high proportion of those classified as low risk or not sexually active at baseline, particularly among men. This highlights the need for continuous engagement of those in transient periods of sexual risk-behaviour in settings with high background HIV prevalence to support their evolving prevention needs. Reaching individuals during periods of risk is also important for the effectiveness and managing costs of prevention services. Self-risk assessment can also be offered for those less likely to attend services.

Reported self-perception of future HIV risk was not useful for identifying those at elevated risk. No incident cases of HSV-2 or HIV occurred among individuals who reported perceived future risk of infection. This was consistent with an earlier study in Manicaland which found low accuracy of HIV risk perception and high HIV incidence in those not perceiving a risk of acquiring HIV, particularly among young people^29^. The utility of a question on future risk perception is likely limited and may miss individuals who could acquire HIV. The poor self-risk perception among youth highlights the utility of a tool to pose non-judgemental questions as part of counselling to aid clients and providers in assessing prevention need. 1% of participants who were HIV-negative and 1/3 who were HIV positive at baseline had a known positive partner, indicating this as an important question to include for prevention referral for those whose partners may not have reached viral suppression.

Previously established risk-behaviours, such as transactional sex and sex between men, were not associated with HIV/HSV-2 acquisition in our study. This should not be interpreted as evidence to remove them from the risk-differentiation tool: the limited sample size and general-population sampling may have limited our ability to detect all associations and assess the predictive capacity of the tool. Some of the follow-up time coincided with COVID-19 lockdowns in Zimbabwe which may have caused a change in sexual risk-behaviour, potentially reducing high risk sexual behaviours and incident infections^30^. Sexual risk behaviours were self-reported and are likely to be under-reported despite ICV methods^22^, although likely to be more commonly reported in this study than in a face-to-face HTC setting. Individuals who declined HTC but consented to provide a DBS were included in the analysis; therefore, respondents who may not attend routine HIV testing services were included. 79% of participants took up HTC in the survey and 75% of those who declined self-reported being HIV-positive and on ART. The proportion of women meeting the criteria for the sex work package is high compared to recent estimates for Zimbabwe, which may stem from the question wording^31^.

## Conclusion

We validated an HIV risk-differentiation tool for the prioritization of prevention services that resulted in 77% sensitivity in predicting HIV/HSV-2 acquisition among young adults in eastern Zimbabwe who had ever had sex. Self-reported questions on risk behaviour were predictive in identifying young individuals who could benefit from HIV prevention services, but missed a large population who reported no sexual activity but subsequently acquired HIV infection after sexual debut. Counselling on sexual risk behaviours and HIV prevention options, particularly among young adults and those recently, or likely to become, sexually active, should be considered for all prevention referrals in settings or populations with high HIV prevalence. This counselling should emphasise what to do if prevention demand does change and support such change. Rapid changes in sexual behaviour over the study period highlights the importance of encouraging regular engagement in youth-friendly HIV prevention and testing services, with opportunities for sexual education promoting HIV prevention prior to sexual debut. While risk-differentiation tools can be useful in supporting HIV prevention demand and linkage to relevant services, the large share of infections occurring among persons classified as ‘low risk’ indicates that responses to a risk-differentiation tool should not be used to restrict access to prevention services for any individuals requesting prevention, even if deemed at low risk. Such tools should be used to support a conversation with clients to support HIV prevention demand and ensure linkage to differentiated services over time as risk changes to meet their needs and reduce the potential for HIV acquisition.

## Data Availability

Due to the sensitive nature of data collected, including information on HIV status, treatment and sexual risk behaviour, the Manicaland Centre for Public Health does not make full analysis datasets publicly available. Summary datasets of household and background sociodemographic individual questionnaire data, covering rounds 1-8 (1998-2021), are publicly available for download via the Manicaland Centre for Public Health website here - http://www.manicalandhivproject.org/data-access.html. Quantitative data used for analyses produced by the Manicaland Centre for Public Health are available on request following completion of a data access request form here - http://www.manicalandhivproject.org/data-access.html. Additionally, summary HIV incidence and mortality data spanning rounds 1-6 (1998-2013), created in collaboration with the ALPHA Network are available via the DataFirst Repository here - https://www.datafirst.uct.ac.za/dataportal/index.php/catalog/ALPHA/about

## Acknowledgements

We are very grateful to the research team working at the Manicaland Centre for Public Health, as well as to the study participants in Manicaland, for their contribution to this study.

SG, SD, DL, SM and LM conceptualised the study. CN, TD, TM, PM, and RM had major roles in collection and management of data used in this study. JM carried out laboratory testing related to the study and aided with interpretation of laboratory results. LM analysed the data. LM, SG, JWI-E, RB, DL and SD contributed to interpretation of results. LM wrote the initial report which was reviewed and revised by all co-authors. All authors had final responsibility for the decision to submit for publication.

