## Supplementary information for "Dynamic HIV risk differentiation among youth: Validation of a tool for prioritization of prevention in East Zimbabwe"

**Supplementary material: Evaluation of a risk differentiation tool to predict HIV and HSV-2 acquisition in 12 months of follow-up among young men and women in east Zimbabwe**

**Contents of supplementary material**

Eligibility criteria for participation in 12 month follow up	2

Figure S1: Flowchart of participation from baseline to 12 month follow up	2

Figure S2: Draft risk differentiation tool for people testing HIV Negative in routine HIV testing services	3

Figure S3: Draft risk differentiation tool and corresponding Manicaland study questions	4

Figure S4: HIV and HSV-2 testing activities according to collection of dried blood spot (DBS) and provider-initiated testing and counselling (PITC)	5

Detection of HIV infection	7

Figure S5: Zimbabwe Ministry of Health and Child Care National HIV Testing Algorithm	8

Detection of HSV-2 infection	9

Figure S6: Definitions of numerator and denominator used to calculate combined HIV and HSV-2 incidence	9

Table S1: Baseline participation rates and 12-month follow up rates by sex	10

Figure S7: Proportions of participants reporting sexual risk behaviours……………………………11

Table S2: HIV, HSV-2 and combined HIV/HSV-2 incidence by 5 year age group	12

**Eligibility criteria for participation in 12 month follow up**

- Control arm for behavioural economics/community psychology interventions
  - Females 15-24 years, HIV-negative (PITC/lab)
  - Males 15-29 years, HIV-negative (PITC/lab)
  - Females 15-24 years, HIV-positive (originally supposed to be 10% on a site-by-site basis but insufficient numbers so all were selected)
  - Males 15-29 years, HIV-positive (originally supposed to be 10% on a site-by-site basis but insufficient numbers so all were selected)
- Intervention arm for behavioural economics/community psychology interventions
  - Females 15-17 years, HIV-negative (PITC/lab)
  - Females 15-24 years, HIV-positive (originally supposed to be 10% on a site-by-site basis but insufficient numbers so selected all)

**Figure S1: Flowchart of participation from baseline to 12 month follow up**

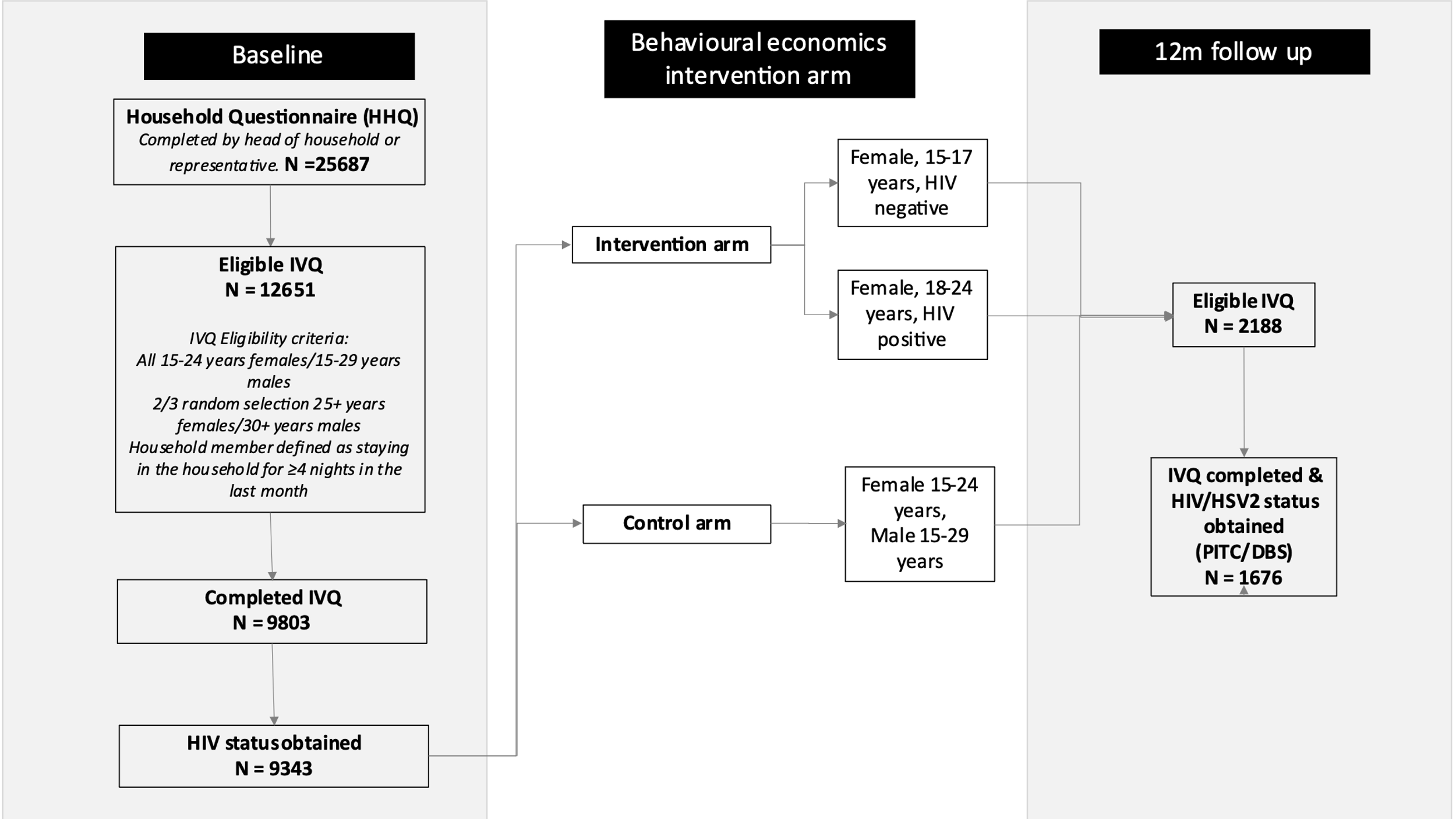

**Figure S2: Draft WHO risk differentiation tool for people that tested HIV negative in routine HIV testing services**

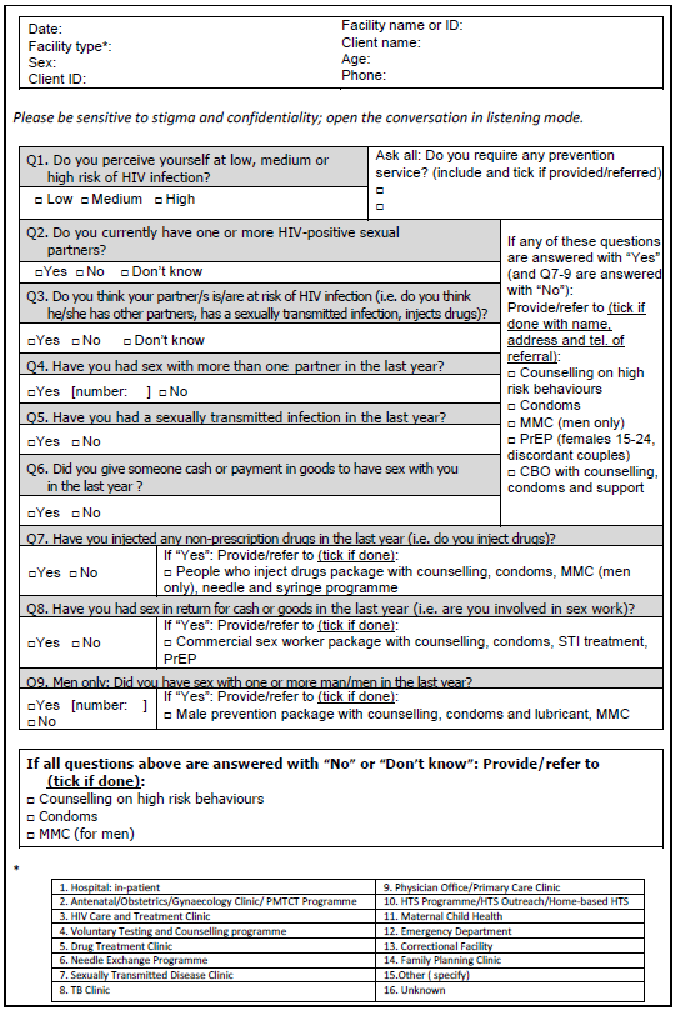

**Figure S3: WHO risk differentiation tool and corresponding Manicaland study questions**

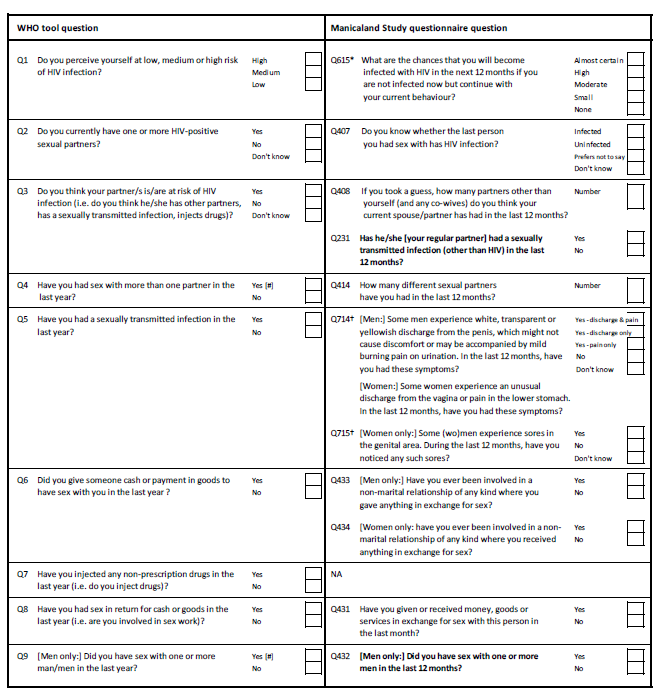

**Figure S4: HIV and HSV-2 testing activities according to collection of Dried Blood Spot (DBS) and Provider Initiated Testing and Counselling (PITC)**

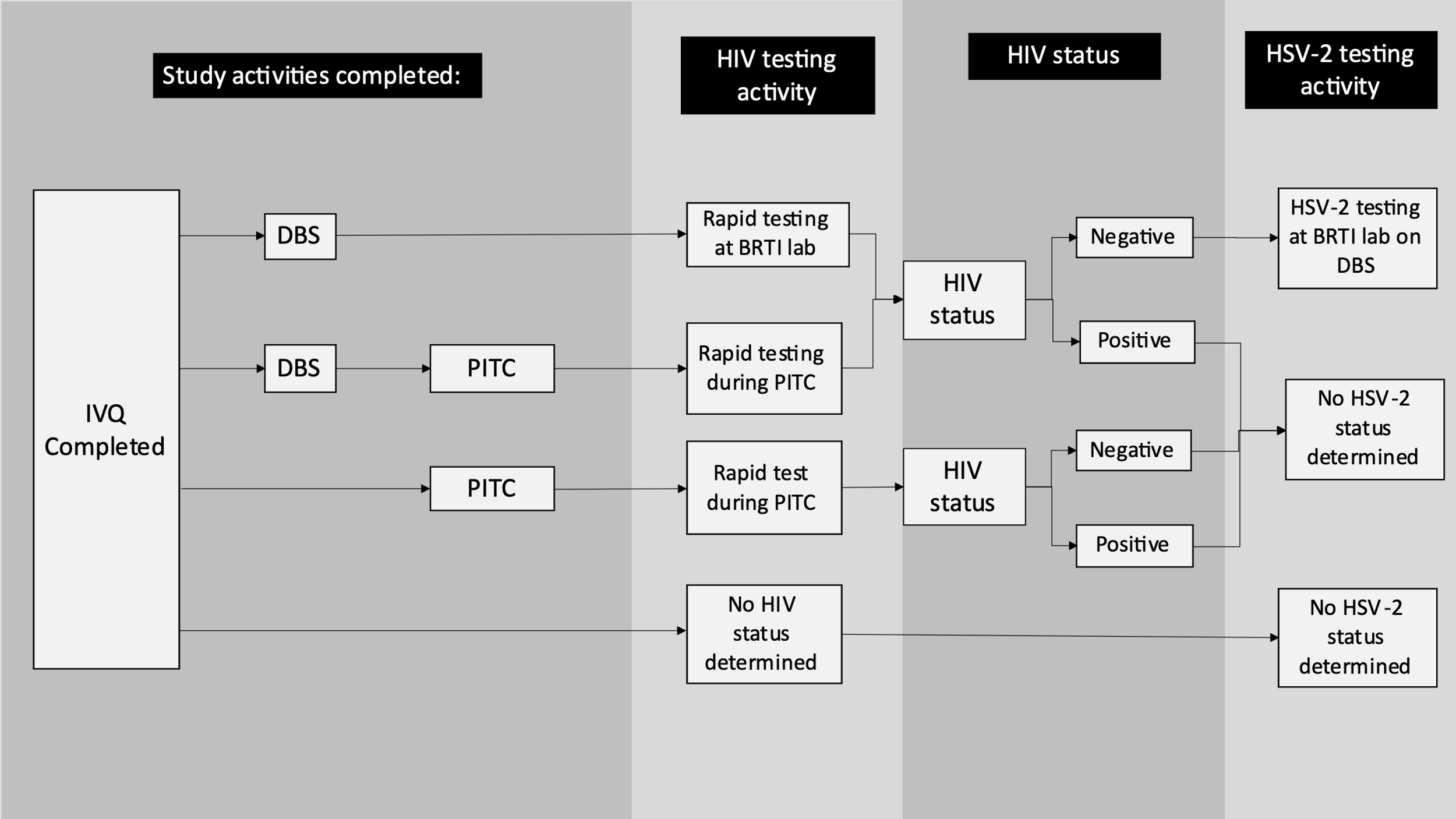

### **Detection of HIV infection**

HIV testing was done by appropriately trained and accredited staff following the standard procedures and testing algorithm used in the ZMoHCC routine PITC services. Currently, the principal ZMoHCC approved algorithm uses Determine as a first screening test, First Response or Chembio to confirm positive results, and INSTI as a tiebreaker). For all individuals declining PITC but wishing to participate in the baseline survey for the research and a random sample 10% sample of other participants, DBS specimens (6 spots total, approximately 75 µL of blood per spot) will be collected onto Whatman filter papers during the baseline survey, barcode-labelled with the participant survey ID, placed in manila envelopes to avoid cross-contamination, transported to the Biomedical Research and Training Institute (BRTI) laboratory in Harare, and tested using the same ZMoHCC algorithm. For long-term storage, DBS cards will be kept at -70^o^C at the BRTI laboratory for specimen testing and archiving.

Quality control (QC) procedures

QC panels consisting of a positive and negative control specimen will be sent to the field staff to ensure test kits are performing correctly. QC panels were performed approximately once a week by each field staff performing HTC and results will be recorded on a QC log form. Additionally, proficiency testing (PT) panels (which included blinded HIV-positive and HIV-negative specimens) were sent to each field staff performing HTC for quality assurance (QA) monthly. One PT panel was prepared for each HTC staff person for each survey round and was administered midway through the course of the survey. Refresher training to ensure tests are performed and interpreted properly and the testing algorithm is being followed correctly were conducted throughout survey implementation. Both QC and PT panels were produced using dried tube specimens which were conducive to field testing conditions.

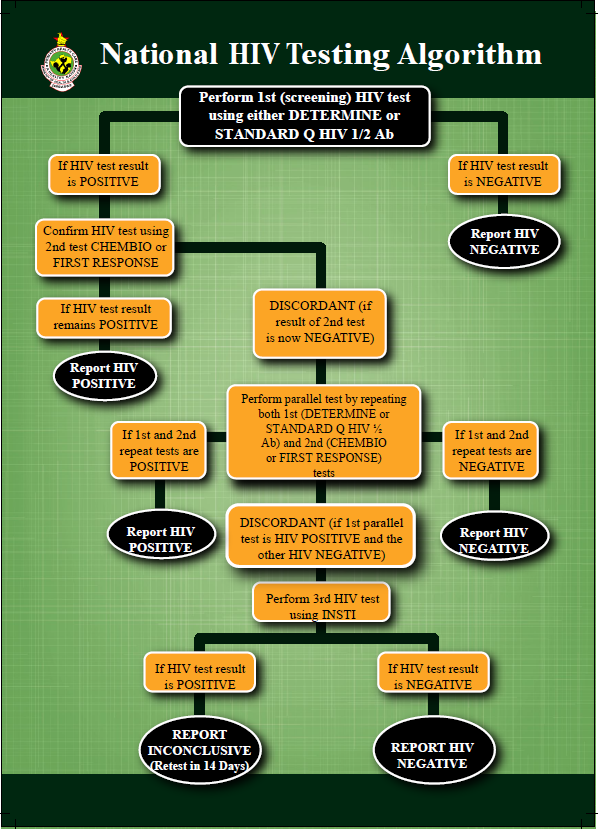
**Figure S5: Zimbabwe Ministry of Health and Child Care National HIV Testing Algorithm**

**Detection of HSV-2 Infection**

For Herpes Simplex virus 2 (HSV-2), DBS specimens were collected onto Whatman 903 filter papers from all baseline survey participants who provide informed consent and at 12 month follow participants randomised to the control arm of all experiments.

From each of the participant's DBS, a 6 mm-diameter disk was punched out from the filter paper into a 96-well microtiter plate and soaked overnight (18-24hours) at 4 °C in 200 µL of phosphate-buffered saline (PBS) (pH 7.4). After the overnight elution step, the eluates were diluted at 1:4 using the kit diluent (HerpeSelect 2 ELISA Immunoglobulin G (IgG) kit (Focus Diagnostics). Each specimen was then tested according to all the other manufacturer’s instructions. Antibodies were qualitatively detected using a HerpeSelect 2 ELISA Immunoglobulin G (IgG) kit (Focus Diagnostics) and the results was be interpreted according to the manufacturer’s instructions. In this assay, type-specific IgG antibodies to HSV-2 were detected in DBS elusion using purified recombinant HSV-2 glycoprotein G antigen (gG2). The principle of the Focus diagnostic HerpeSelect IgG2 assay is as follows: polystyrene micro-wells were coated with recombinant gG2 protein antigen; diluted serum samples and controls were incubated in the wells to allow specific antibody present in samples to react with the antigen; non-specific reactants were removed by washing and peroxidase conjugated anti-human IgG was added that reacts with specific IgG; excess conjugate was removed by another washing step; enzyme substrate and Chromogen were added and the colour change was allowed to develop; after adding a stop reagent, the resultant colour change was quantified by a spectrophotometric reading of Optical Density (OD) (at wavelength 450 nm); and sample OD readings were then compared with reference cut-off OD readings to determine positive and negative results.

For quality control, each plate run (wells or trips) included the cut-off calibrator and all 3 controls. All specimens, controls and the cut-off calibrators were run in single wells were used. The cut-off calibrator was run in triplicate. The tests included a minimum of one blank well (containing sample diluent only) for instrument calibration purposes. The cut-off calibrator was formulated to give the optimum differentiation between negative and positive sera. The mean value for the cut-off calibrator must be within 0.100-0.700 OD units. All replicates cut-off calibrators’ ODs should be within 0.10 absorbance units from the mean value. The results were reported as index values relative to the cut-off calibrator. To calculate index values, the specimen OD values were divided by the mean of the cut-off calibrator absorbance values.

1. The high positive control index value should be >3.5
2. The low positive control index value should be >1.1 and <3.5
3. The negative control index value should be <0.9

If the cut-off calibrator or controls were not within these parameters, patients test results were considered to be equivocal and the assay was repeated. The positive and negative controls were intended to monitor for substantial reagent failure; the positive control was not used as an indicator for cut-off value precision.

In individuals whose samples went from baseline negative to 12 month follow up equivocal, the testing process was repeated in full for the 12 month follow up samples using a separate DBS punch. In individuals whose samples went from baseline negative to 12 month follow up positive, the testing process was repeated in full for the baseline and 12 month follow up samples using a separate DBS punch.

**Figure S6: Definitions of numerator and denominator used to calculate combined HIV/HSV-2 incidence**

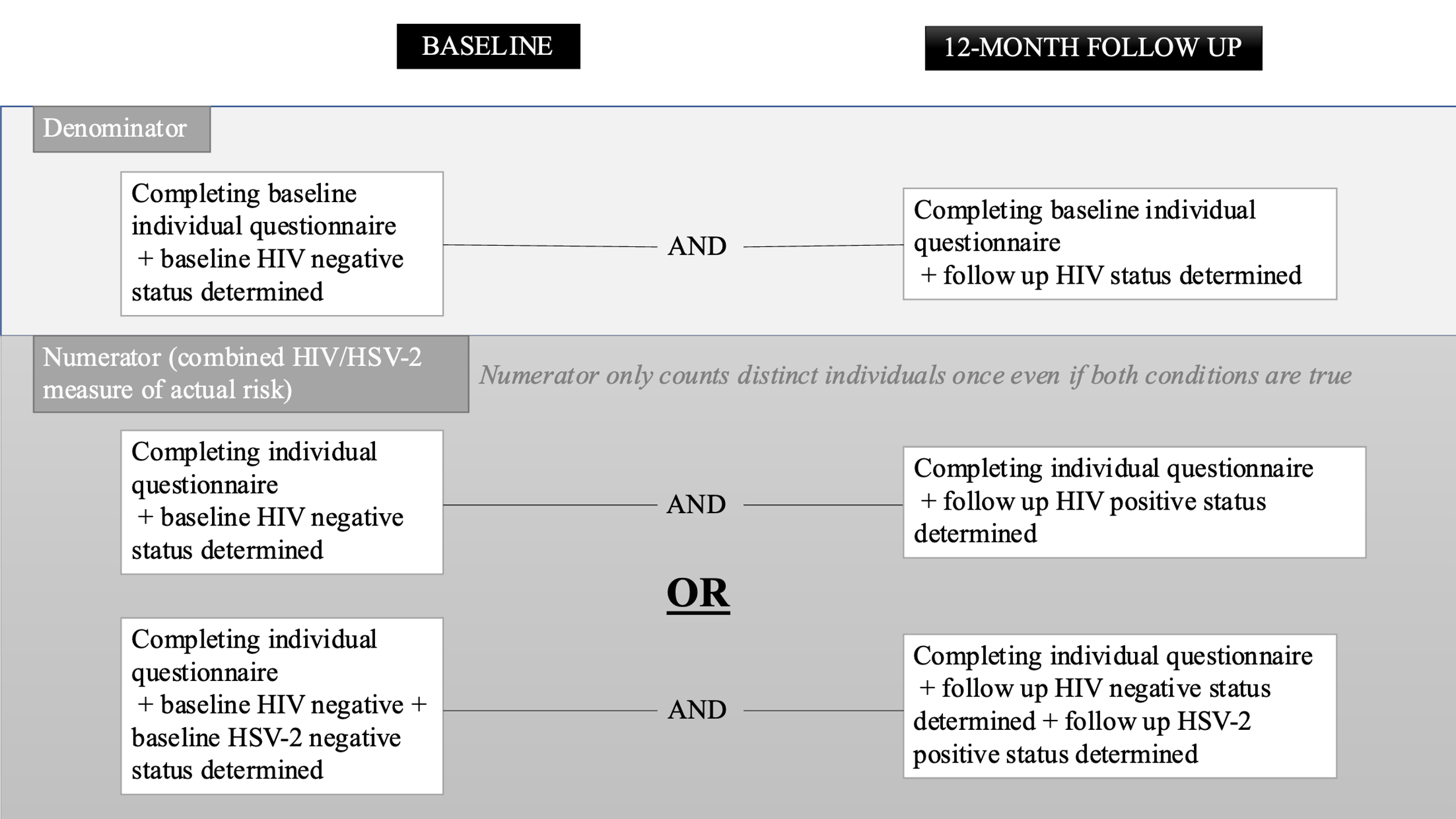

**Table S1: Baseline participation rates and 12-month follow up rates by sex**

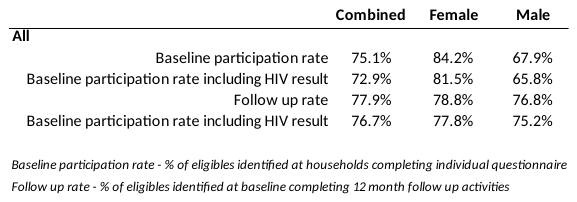

**Figure S7 – Proportions of HIV negative young women (n=957) and men (n=675) at baseline reporting to have not started sexual activity, sexually active but no risk behaviour, or sexually active with risk behaviour in the last 12 months (≥1 of having a known HIV-positive partner, having multiple sex partners in the past year, perception of partner risk, having had a sexually transmitted infection in the past year, transactional sex, sex between men, and injecting drug use)**

%

|  | Women 15-24 yrs | Men 15-29 yrs |
| --- | --- | --- |
| Not sexually active | 57.5758 | 61.2184 |
| No risk behaviour reported | 25.3918 | 10.847 |
| Risk behaviour reported | 17.0324 | 27.9346 |

**Table S2: HIV, HSV-2 and combined HIV/HSV-2 incidence by 5-year age group**

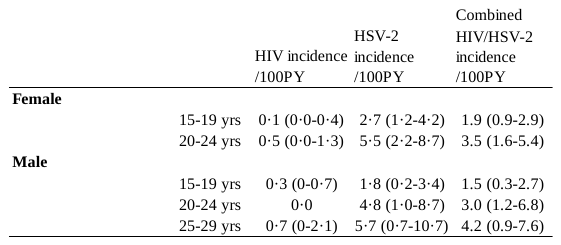
